# Burden of lower extremity peripheral arterial disease among people aged 40 years and older in the Western Pacific region, 1990-2021: a comparative analysis of China, South Korea, Japan, and the Philippines

**DOI:** 10.1101/2025.09.07.25335289

**Authors:** Dalong Zhao, Cuiping Jiang, Zhiqing Li, Yuyang Wang, Weifeng Xie, Jun Chen

**Author notes:** Corresponding author: Jun Chen (JC).

## Abstract

**Background:** Lower extremity peripheral arterial disease (PAD) has a significant adverse impact on lower limb function and the quality of daily life, making it an important public health issue. PAD is closely associated with cardiovascular diseases. However, compared to conditions such as myocardial infarction and stroke, research on PAD remains insufficient and has not received adequate attention in global health policies. The Global Burden of Disease (GBD) study aims to integrate all available epidemiological data through a unified measurement framework, standardized estimation methods, and transparent data sources, enabling comparisons of disease burden across time, sex, age, and countries. This study utilizes data from GBD 2021 to explore the epidemiological trends of PAD among people aged 40 years and older in China, Japan, South Korea, and the Philippines from 1990 to 2021, highlighting regional differences.

**Methods:** This study analyzed the burden of PAD among individuals aged 40 years and older in four countries in the Western Pacific region, using publicly available data from GBD 2021, including prevalence, incidence, and disability-adjusted life years (DALYs). The 95% uncertainty intervals (95% UI) reflect the uncertainty associated with each specific indicator by country, age, sex, and year. Age-standardized rates (ASRs) were calculated to enable fair cross-country comparisons. We used the Socio-Demographic Index (SDI) to quantify the developmental status of each country, which is a composite metric derived from indicators of income per capita, educational attainment, and fertility. Joinpoint regression analysis was employed to assess temporal trends. The Bayesian age-period-cohort (BAPC) model was used to predict the incidence until 2041.

**Results:** In 2021, the Philippines had the highest incidence, prevalence, and DALY rates, followed by China and Japan, with South Korea having the lowest. The affected population in the Philippines also tended to be younger, with China being the next youngest. China had the highest number of PAD (Peripheral Artery Disease) patients, with 2,447,372 new cases in 2021 and 28,474,886 prevalent cases. Among the four countries, the disease burden was significantly higher in women than in men. Incidence, prevalence, and DALY rates all peaked in older age groups. From 1990 to 2021, the burden in China and the Philippines showed an upward trend, while that in Japan and South Korea exhibited a downward trend. Future projections indicate that the burden in China will increase slightly, the incidence in Japan and South Korea will continue to decline, and the burden in the Philippines will remain relatively stable.

**Conclusions:** The burden of PAD is relatively heavy in the Western Pacific region. Countries with lower SDI need to address its social and economic impacts, and develop targeted strategies for optimal prevention and treatment, taking into account factors such as economy, demographic structure, universal health coverage, and regional distribution. Countries with higher SDI need to focus on disease control for the elderly.

## 1. Introduction

Non-communicable diseases (NCDs) are currently the leading causes of morbidity and mortality worldwide[1,2]. The burden of NCDs is rapidly increasing due to population aging and greater exposure to risk factors for chronic diseases, such as smoking, alcohol consumption, hypertension, diabetes, obesity, and hypercholesterolemia. A quarter of global deaths from NCDs occur in the Western Pacific region[3]. Based on the widely recognized epidemiological transitions across different regions of the world, assessing the current burden of major NCDs at the global or regional level may help inform the development of effective and cost-efficient response strategies. However, during the research process, some diseases are often overlooked. PAD is one of them. It is the third most prevalent atherosclerotic vascular disease, following coronary heart disease and stroke[1]. Its pathological hallmark is the narrowing or occlusion of the arterial lumen in the lower limbs[4]. The clinical manifestations of PAD mainly include intermittent claudication, leg pain, and pain at rest[5], with severe cases potentially leading to limb ischemia and necrosis[6,7]. Although PAD significantly affects the elderly population, epidemiological data indicate a trend toward younger age of onset[2].

The Western Pacific Region is facing multiple public health challenges, including population aging, the rise of cardiovascular risk factors, lifestyle changes associated with socioeconomic development, and the uneven spatial distribution of medical resources[8]. The above factors have made this region a key area for epidemiological studies of PAD. It is also true that PAD has caused a large number of affected groups and disability-adjusted life years in this region over the past three decades. Accurate epidemiological data are the basis for early medical intervention, and are of great practical significance for the development of disease prevention strategies, the optimization of medical resource allocation, and the determination of public health priorities.

Although previous studies have partially explored the prevalence and disease characteristics of PAD on a global or regional scale, these investigations are limited by fragmented content and outdated data. To overcome these limitations, this study leverages the latest data from GBD 2021. In addition to conducting descriptive analyses, it further employs advanced statistical methods such as joinpoint regression analysis and Bayesian age-period-cohort models to deeply investigate the trends in PAD disease burden. The study systematically analyzes the age-sex-time distribution patterns of PAD disease burden and, for the first time, predicts the future disease burden of PAD. Moreover, by utilizing age-standardized indicators, this research quantitatively assesses regional disparities in the PAD disease burden across the Western Pacific region for the first time.

This study focuses on China, Japan, South Korea, and the Philippines, which represent diverse economic, social, and healthcare contexts in the Western Pacific region. We analyzed the prevalence, incidence, and DALYs of PAD from 1990 to 2021. The use of age-standardized rates ensured fair comparisons, Joinpoint regression was employed to identify temporal trends, and the Bayesian age-period-cohort model was used to project the burden up to the year 2041.

## 2. Methods

### Overview of the Study

Publicly available data from GBD 2021 were used for secondary analysis. GBD 2021 is a large-scale global collaborative research initiative aimed at comprehensively quantifying and analyzing the burden and distribution of major diseases, injuries, and risk factors worldwide. GBD 2021 was published by the Institute for Health Metrics and Evaluation (IHME) at the University of Washington, USA. Its data were collected from multiple sources, including vital registration systems, censuses, household surveys, disease-specific registries, and health service contact data. Detailed information on the complete data sources can be accessed online through the GBD 2021 Source Tool on the IHME website (https://ghdx.healthdata.org/gbd-2021/sources). After data collection, DisMod-MR 2.1 — a standardized statistical modeling tool — was used to assess and adjust for potential biases in each dataset. GBD 2021 provides estimates and analyses for 371 diseases and injuries and 88 risk factors across 204 countries and territories globally, stratified by year (from 1990 to 2021), age (from birth to 95 years and older), and sex (male, female, and both sexes). It offers annual data at the global, regional, and national levels from 1990 to 2021. For this study, PAD data from China, South Korea, Japan, and the Philippines for the period 1990–2021 were extracted from GBD 2021. The data were adjusted for country-specific factors such as differences in healthcare infrastructure and diagnostic practices, in order to more accurately reflect the true disease burden in each respective country.

Sociodemographic Index (SDI) is a composite indicator of the development status of a country or region, used to analyze the relationship between health and socioeconomic development. The SDI is derived from a comprehensive assessment of data such as fertility rates among people under 25, mean educational attainment among those aged 15 and older, and income per capita. Its value ranges from 0 to 1, with higher values indicating better development status. Based on SDI values, 204 countries and territories are divided into five distinct tiers: low sociodemographic development, low-middle sociodemographic development, middle sociodemographic development, high-middle sociodemographic development, and high sociodemographic development. Japan and South Korea fall into the high-middle category, while China and the Philippines are classified under the middle sociodemographic development level (Table 1).

**Table 1.** SDI values for four countries, GBD 2019.

| Location | Socio-Demographic Index values (GBD 2019) |
| --- | --- |
| Japan | 0.87 |
| South Korea | 0.878 |
| China | 0.686 |
| Philippines | 0.623 |

### 2.1 Data sources

The prevalence, incidence, and DALYs of PAD in China, South Korea, Japan, and the Philippines, along with their 95% uncertainty intervals, are all available on GBD 2021 (https://vizhub.healthdata.org/gbd-results/).

Only publicly available aggregated data from the GBD 2021 were used, with no access to individually identifiable information.

### 2.2 Burden Description

This study uses incidence, prevalence, and DALYs as core indicators to describe the burden of PAD. Incidence reflects the frequency of new cases of the disease, prevalence indicates the proportion of the population affected by the disease at a certain point in time or over a period, while DALYs represent the sum of years of life lost (YLLs) due to premature death and years lived with disability (YLDs) due to health loss caused by the disease, providing a comprehensive measure of the health impact of the disease. YLLs are calculated by multiplying the number of deaths for each cause-age-sex-region-year by the standard life expectancy at each age, and YLDs are calculated as the product of the estimated age-sex-region-year-specific prevalence and the corresponding disability weights.

Estimates for all measures are reported as 95%UI, which are taken at the 2.5th and 97.5th percentiles of the distributions derived from 100 samples of uncertainty. The 95% UI can reflect the stability and reliability of the data.

Age-standardized rates (per 100,000 population) for each indicator were calculated using the GBD standard population. Age-standardized rates eliminate the influence of differences in age structure among populations on the indicators, ensuring the comparability of disease burden data across different countries and enhancing the validity of disease burden assessment results.

### 2.3 Joinpoint regression analysis

The Joinpoint regression model consists of multiple linear statistical models to analyze the time trend of disease burden[9]. The model takes trend data and fits the simplest joinpoint model allowed by the data. The model starts with the minimum number of connection points and tests if more are statistically significant and added to the model up to the maximum number. The tests of significance use a Monte Carlo Permutation method. Joinpoint regression models capture trend turning points in time series through piecewise linear models to quantify the magnitude of change at each stage. It provides a rigorous statistical framework for the study.

In this study, the Joinpoint regression model was used to conduct segmented regression analyses on the temporal trends of incidence, prevalence, and DALYs. The annual percent change (APC) and average annual percent change (AAPC), along with their 95% uncertainty intervals, were calculated. APC represents the average annual rate of change in disease burden during a specific time period, whereas AAPC reflects the overall average annual rate of change over the entire study period. Whether the observed trend changes were statistically significant was determined by comparing them to zero. A 95% UI that lies entirely above zero indicates a statistically significant upward trend; if it lies entirely below zero, it indicates a statistically significant downward trend; if it includes zero, the trend during that period is not statistically significant. A P-value less than 0.05 was considered statistically significant.

### 2.4 Bayesian age-period-cohort analysis

Bayesian age-period-cohort model was used to predict the future incidence of PAD. The Bayesian age-period-cohort model is a Bayesian extension of the traditional age-period-cohort (APC) model. It addresses the identification challenges inherent in the traditional model and provides more flexible parameter estimation along with better quantification of uncertainty. Its core lies in constraining the parameter space through prior distributions and using posterior inference to quantify the uncertainty of the effects. To ensure the accuracy and robustness of the predictions, we utilize PAD incidence data from 1990 to 2021 to forecast the incidence from 2022 to 2041.

All analyses and visualizations in this study were performed in R 4.5.1(The R Foundation for Statistical Computing, Vienna, Austria).

## 3. Results

### 3.1 Burden of PAD in China, Japan, South Korea, and the Philippines in 2021

The number of incident cases of PAD in China was 28,474,886 (95% UI: 24,446,553–33,220,180), the number of prevalent cases was 24,473,726 (95% UI: 21,099,910–28,542,910), and the DALYs were 171,757 (95% UI: 99,158–301,528). The age-standardized incidence rate (ASIR), age-standardized prevalence rate (ASPR), and age-standardized DALYs were 112.66 (95% UI: 97.75–130.73), 1,331.13 (95% UI: 1,147.49–1,544.17), and 8.36 (95% UI: 4.87–14.33), respectively. In Japan, the number of incident cases of PAD was 376,222 (95% UI: 323,285–432,518), the number of prevalent cases was 4,885,245 (95% UI: 4,233,083–5,631,107), and the DALYs were 45,590 (95% UI: 33,650–63,004). The ASIR, ASPR, and age-standardized DALYs were 113.02 (95% UI: 97.48–130.17), 1,309.38 (95% UI: 1,135.62–1,515.95), and 9.66 (95% UI: 7.00–14.13), respectively. In South Korea, the number of incident cases was 95,649 (95% UI: 82,020–110,495), the number of prevalent cases was 1,070,954 (95% UI: 935,686–1,245,048), and the DALYs were 7,482 (95% UI: 5,182–11,324). The ASIR, ASPR, and age-standardized DALYs were 100.88 (95% UI: 86.62–116.11), 1,132.29 (95% UI: 989.94–1,314.26), and 7.94 (95% UI: 5.55–11.89), respectively. In the Philippines, the number of incident cases was 104,168 (95% UI: 89,923–121,956), the number of prevalent cases was 1,095,304 (95% UI: 940,456–1,279,930), and the DALYs were 8,635 (95% UI: 5,679–13,901). The ASIR, ASPR, and age-standardized DALYs were 123.84 (95% UI: 106.98–144.23), 1,398.11 (95% UI: 1,209.27–1,619.81), and 11.82 (95% UI: 7.55–19.42), respectively (Table 2).

**Table 2.**
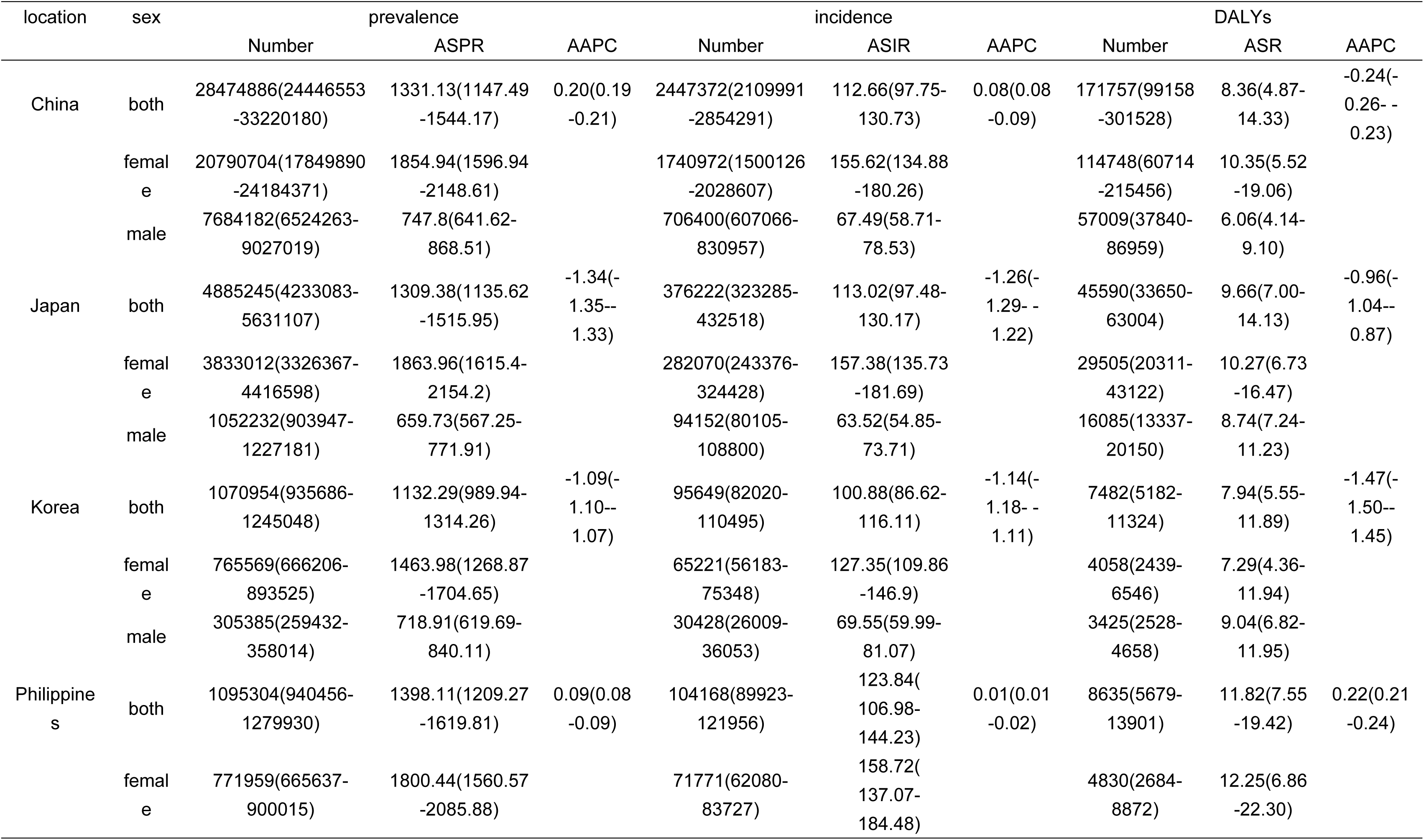
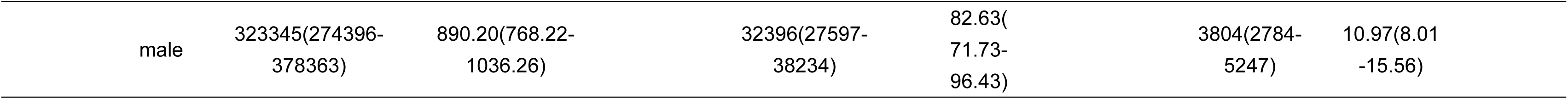
Burden of PAD among China, Japan, South Korea and Philippines in 2021 and AAPC from 1990 to 2021.

Among different age groups, the Philippines and Japan exhibited the highest incidence rates in the 75–79 age group, at 817.39 (95% UI: 544.45–1152.27) and 880.10 (95% UI: 591.12–1243.70), respectively. South Korea and China had the highest incidence rates in the over-80 age group, at 845.30 (95% UI: 625.91–1093.14) and 704.91 (95% UI: 522.57–910.88), respectively. Across all four countries, the highest prevalence and DALYs rates were observed in the over-80 age group. Among all age groups, females bore a heavier burden of lower extremity peripheral artery disease (Figs 1, 2, 3, and 4).

**Fig 1.**
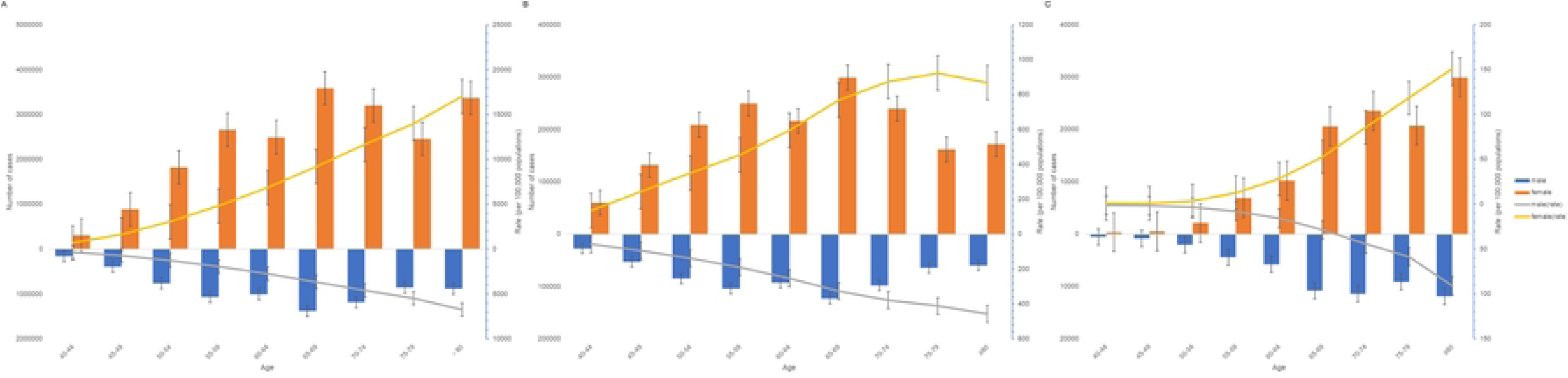
Burden of PAD regarding the sex and age in China in 2021. A Prevalence, B Incidence, C DALYs.

**Fig 2.**
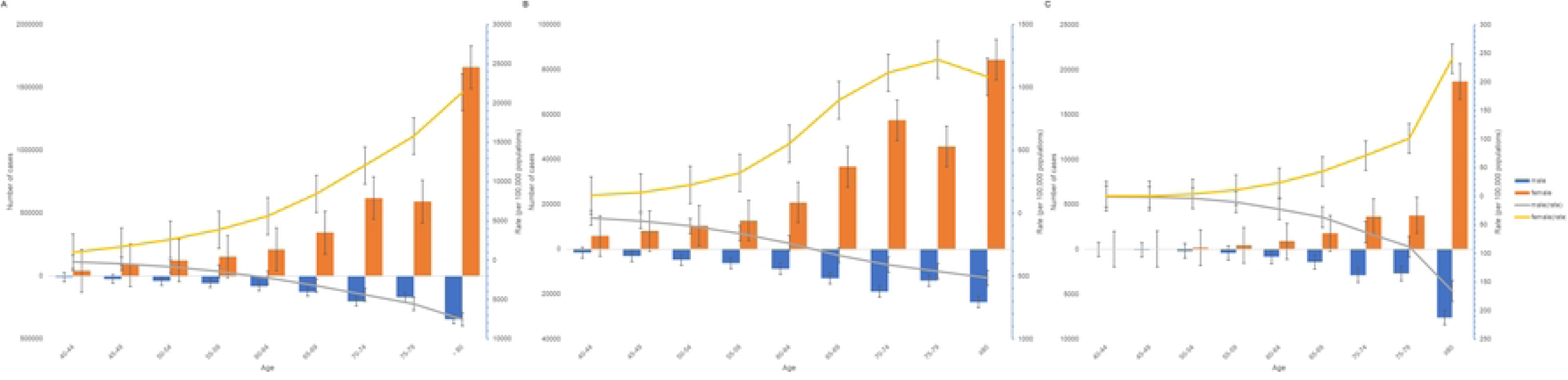
Burden of PAD regarding the sex and age in Japan in 2021. A Prevalence, B Incidence, C DALYs.

**Fig 3.**
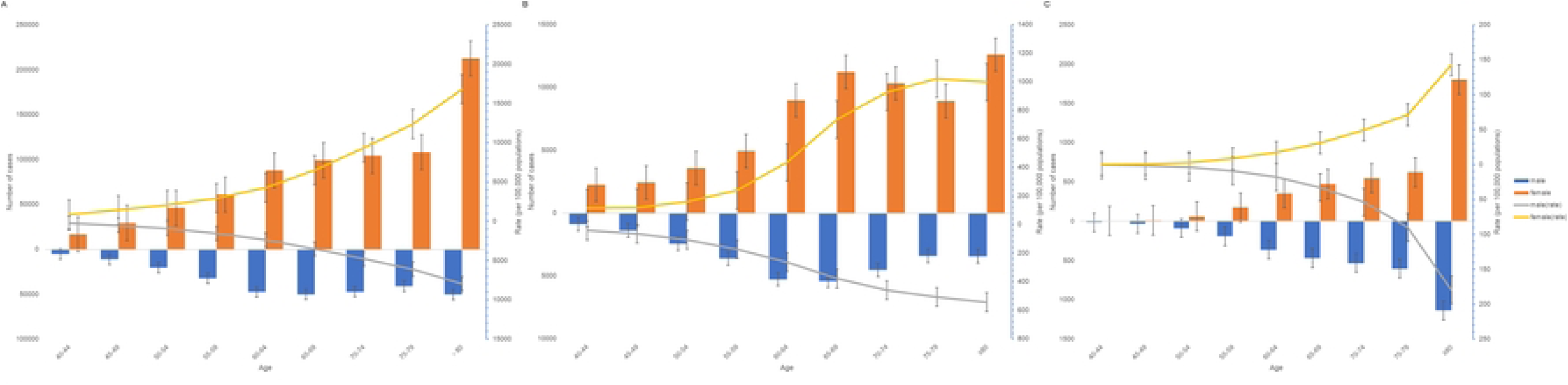
Burden of PAD regarding the sex and age in South Korea in 2021. A Prevalence, B Incidence, C DALYs.

**Fig 4.**
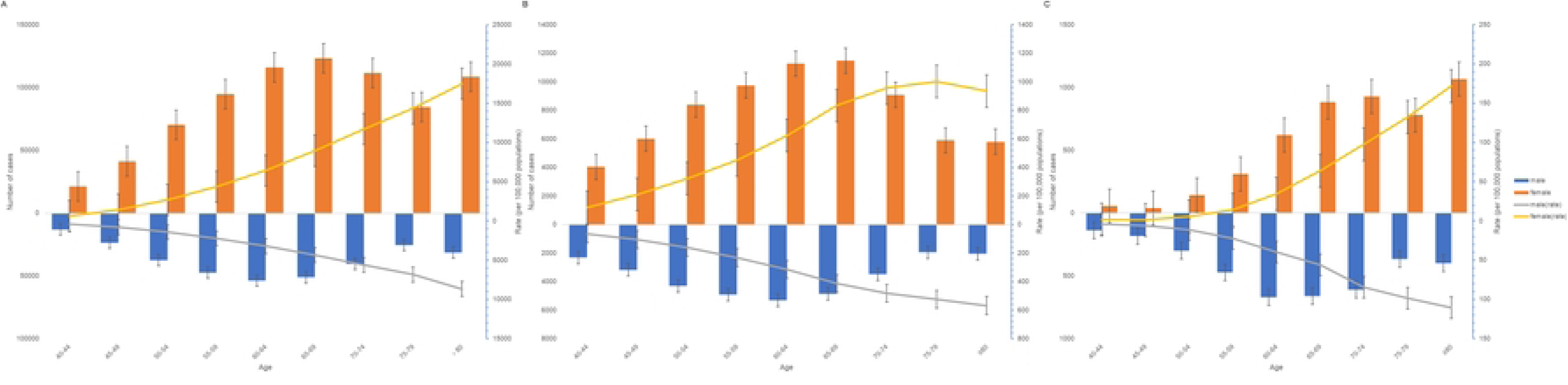
Burden of PAD regarding the sex and age in Philippines in 2021. A Prevalence, B Incidence, C DALYs.

### 3.2 Trends in PAD in China, Japan, South Korea, and the Philippines from 1990 to 2021

Between 1990 and 2021, the burden of PAD varied among the four countries. China and the Philippines experienced an increasing trend in PAD burden, while Japan and South Korea saw a continuous decline. In the Philippines, the AAPC for age-standardized DALY rate of PAD was 0.22 (95% UI: 0.21–0.24), the AAPC for ASPR was 0.09 (95% UI: 0.08–0.09), and the AAPC for ASIR was 0.01 (95% UI: 0.01–0.02). In China, although the disability rate due to PAD decreased, the AAPC for age-standardized DALY rate was -0.24 (95% UI: -0.26 – -0.23); however, both ASPR and ASIR showed an upward trend, with AAPC of 0.20 (95% UI: 0.19–0.21) and 0.08 (95% UI: 0.08–0.09), respectively. In contrast, both Japan and South Korea witnessed a significant decline in PAD burden. In Japan, the AAPC for age-standardized DALY rate was -0.96 (95% UI: -1.04 – -0.87), and the AAPCs for ASPR and ASIR were -1.34 (95% UI: -1.35 – -1.33) and -1.26 (95% UI: -1.29 – -1.22), respectively. In South Korea, the AAPC for age-standardized DALY rate was -1.47 (95% UI: -1.50 – -1.45), and the AAPCs for ASPR and ASIR were -1.09 (95% UI: -1.10 – -1.07) and -1.14 (95% UI: - 1.18 – -1.11), respectively (Fig 5).

**Fig 5.**
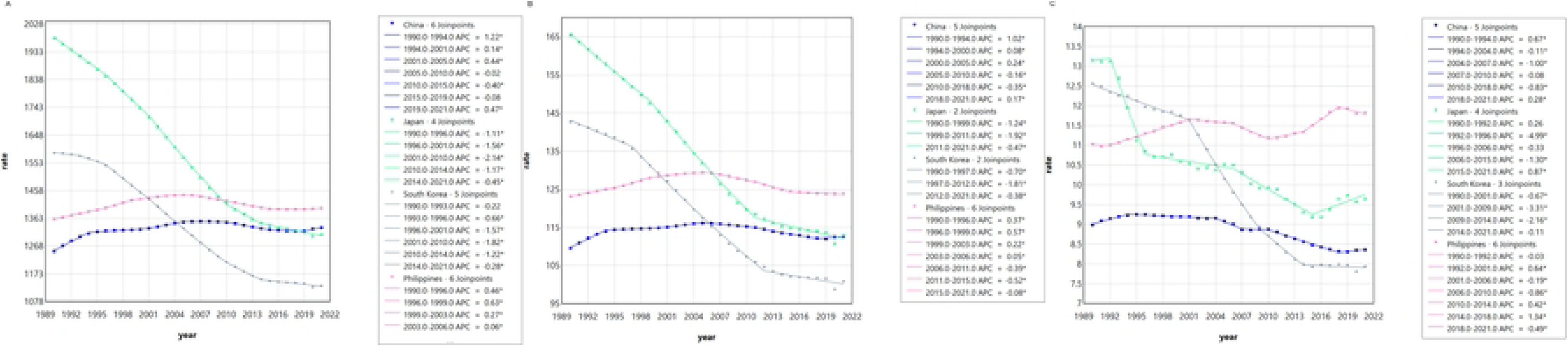
APC among four countries from 1990 to 2021. A Prevalence, B Incidence, C DALYs.

### 3.3 Forecast of the PAD Burden in China, South Korea, Japan, and the Philippines from 2022 to 2041

Bayesian age-period-cohort model was used to predict the burden of PAD in these four countries from 2022 to 2041. In China, the future PAD burden is expected to show a slight increase. The age-standardized incidence rate (ASIR) of PAD in China will be 112.70 (95% UI: 111.67–113.73) in 2022 and 114.10 (95% UI: 92.49–137.11) in 2041. Among them, females represent the primary affected group, with the ASIR for women being 155.88 (95% UI: 154.00–157.75) in 2022 and 161.61 (95% UI: 118.41–208.48) in 2041. In contrast, the incidence among Chinese males shows a downward trend, decreasing from 67.30 (95% UI: 66.24–68.33) in 2022 to 63.52 (95% UI: 47.04–79.89) in 2041 (Fig 6). Both Japan and South Korea are projected to experience a significant decline in the future PAD burden. Japan’s ASIR is estimated to be 111.21 (95% UI: 103.86–118.52) in 2022 and 89.63 (95% UI: 36.19–160.93) in 2041 (Fig 7). South Korea’s ASIR is projected to decrease from 99.34 (95% UI: 93.02–105.66) in 2022 to 79.95 (95% UI: 34.52–141.51) in 2041 (Fig 8). In the Philippines, the future PAD burden is expected to remain stable. The ASIR in the Philippines is forecasted to be 123.72 (95% UI: 122.68–124.81) in 2022 and 122.00 (95% UI: 102.15–143.30) in 2041 (Fig 9).

**Fig 6.**
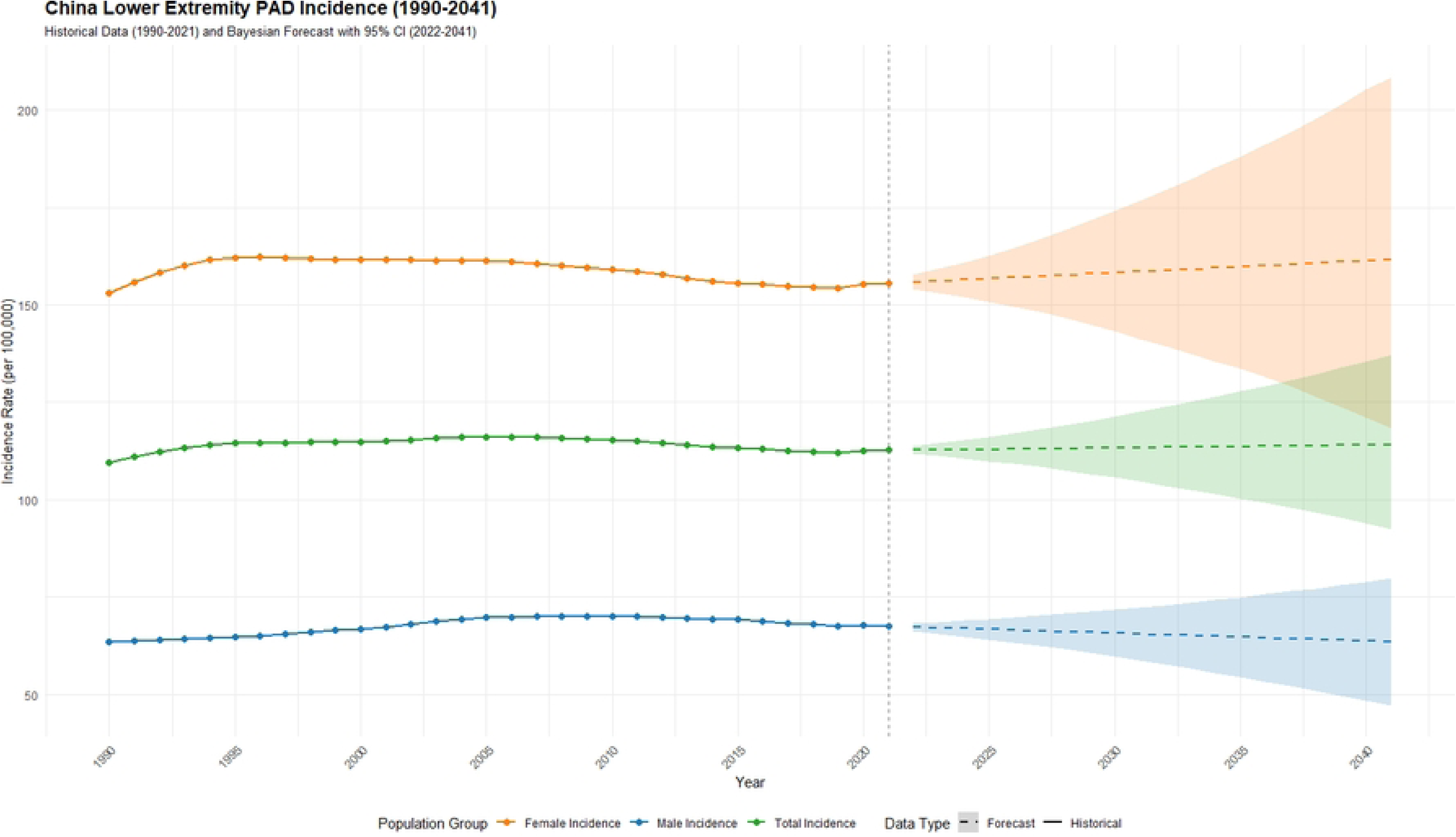
Predicted age-standardized incidence rates of PAD regarding the sex in China from 2022 to 2041.

**Fig 7.**
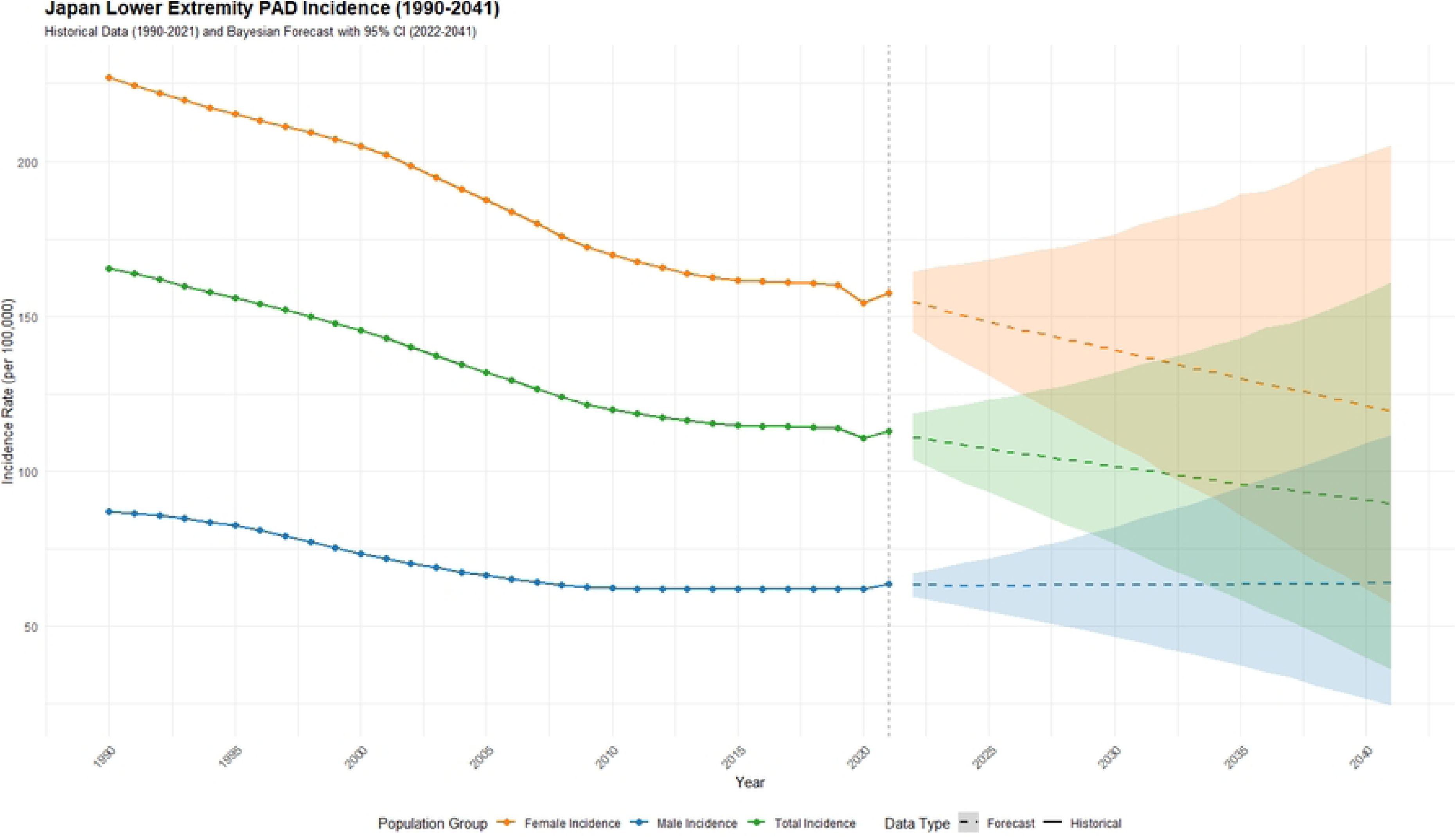
Predicted age-standardized incidence rates of PAD regarding the sex in Japan from 2022 to 2041.

**Fig 8.**
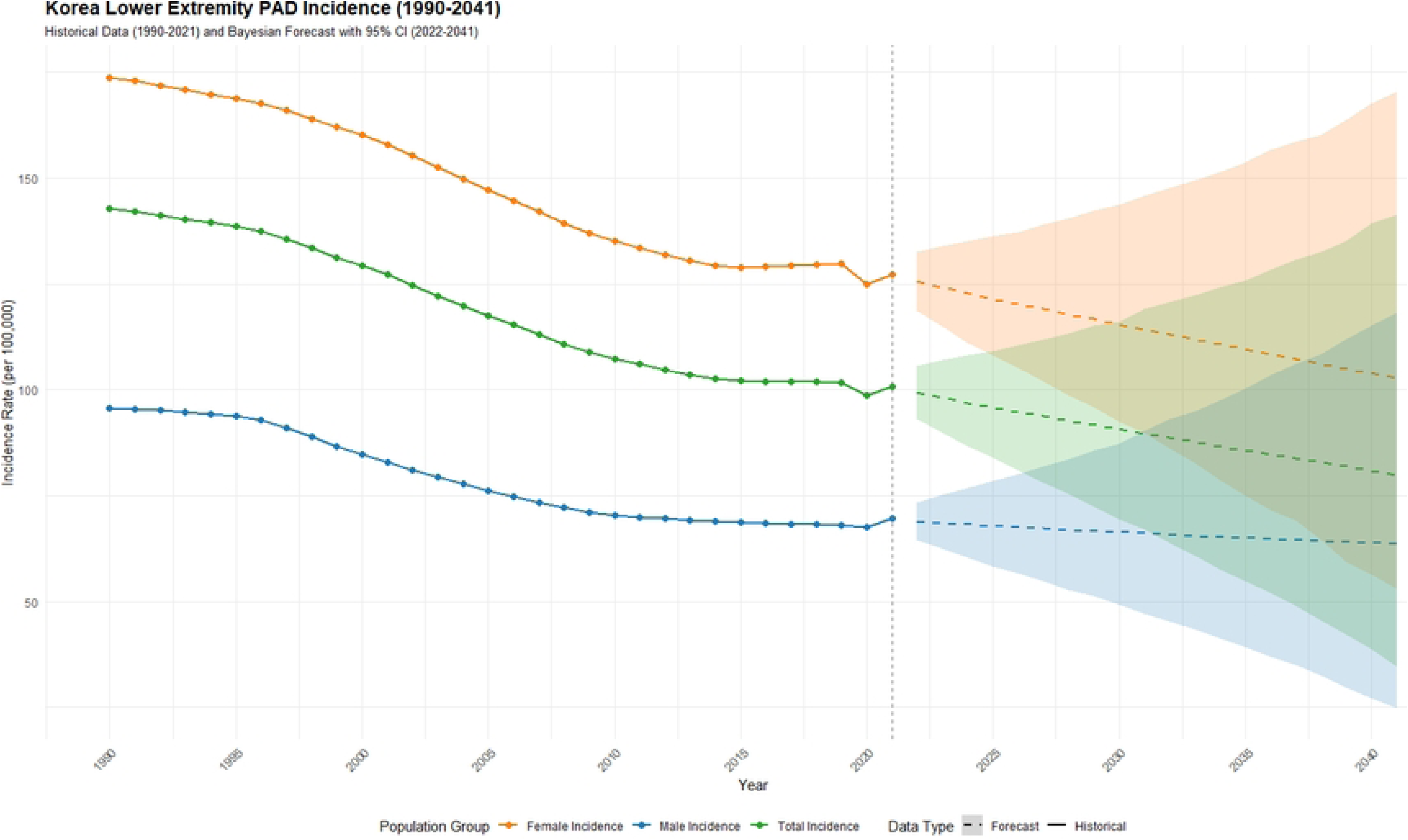
Predicted age-standardized incidence rates of PAD regarding the sex in South Korea from 2022 to 2041.

**Fig 9.**
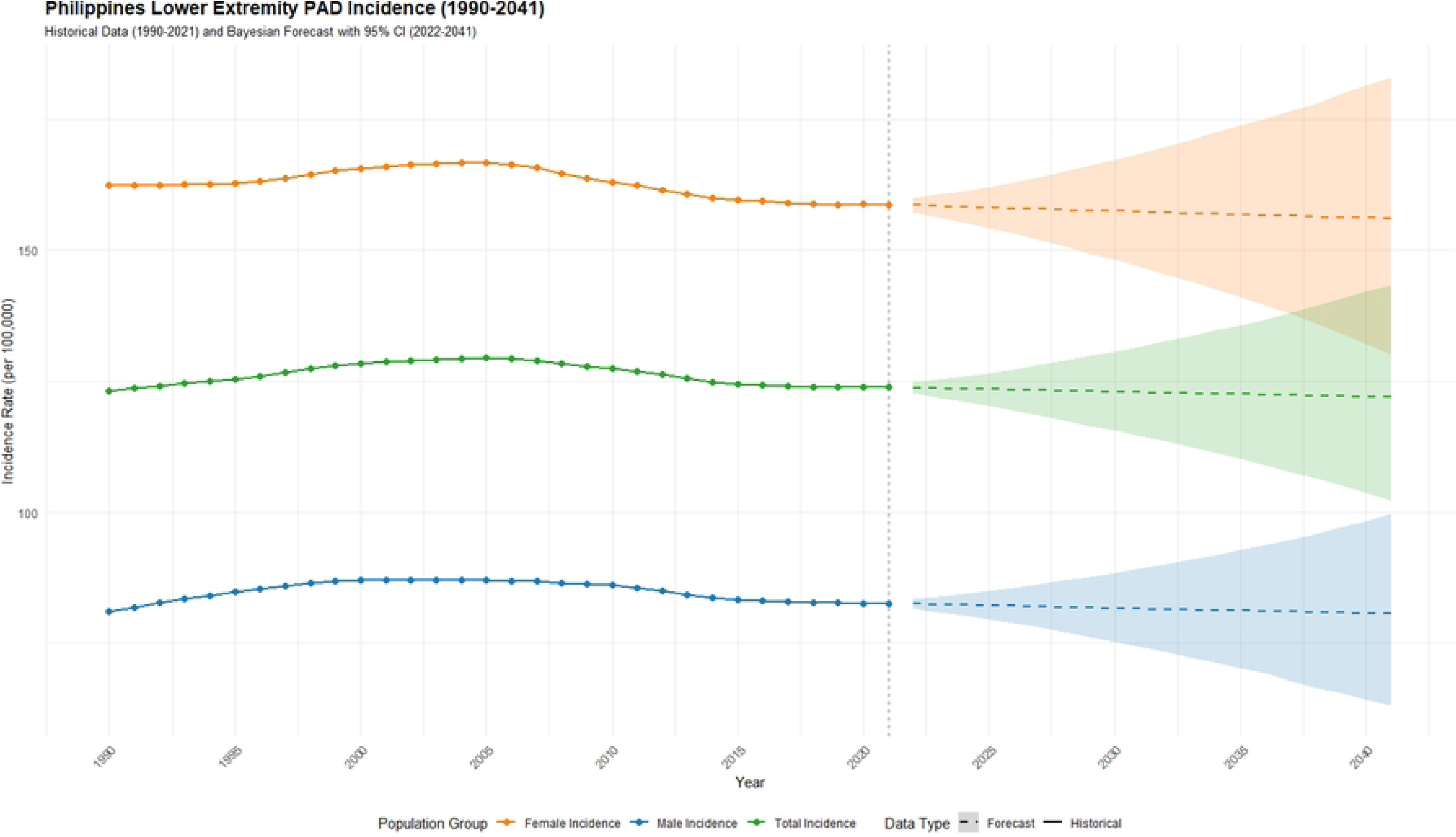
Predicted age-standardized incidence rates of PAD regarding the sex in Philippines from 2022 to 2041.

## 4. Discussion

This study aims to describe and analyze the burden of PAD in China, Japan, South Korea, and the Philippines, revealing differences in trends, age, and gender across regions with varying economic and cultural backgrounds in the Western Pacific. In China, the incidence and prevalence of PAD have shown a slight increase. Japan and South Korea have experienced a significant decline in the PAD burden. The Philippines has relatively stable trends in both incidence and prevalence, but there has been a notable increase in disability burden. Compared to the other three countries, the age structure of the PAD-affected population in the Philippines is younger, and the disability burden also tends to be more concentrated among younger individuals. Across all countries, the PAD burden is significantly higher among women than men.

PAD has now become a major global health challenge. Worldwide, the number of adults affected by this condition has exceeded 230 million[10]. Among atherosclerotic vascular diseases, PAD ranks as the third most prevalent, following only coronary artery disease and stroke. Over the past three decades, in the Western Pacific region, PAD has emerged as one of the key drivers contributing to the increasing burden of cardiovascular disease and rising mortality rates. Patients with PAD face a potential risk of lower limb amputation, with those suffering from severe ischemic necrosis or concurrent infection being at significantly higher risk of amputation[11]. Amputation can lead to a range of negative consequences, including impairments in motor function, mental health disorders, reduced social participation, and increased medical and economic burdens[12]. However, existing research exhibits significant limitations: on the one hand, there is a lack of data on PAD incidence trends, and on the other, there is insufficient evidence regarding regional disparities. The current cardiovascular disease epidemiological assessment system demonstrates a notable bias in focus, with research resources disproportionately concentrated on coronary artery disease and stroke, while long-term neglect has characterized the evaluation of the disease burden associated with lower extremity peripheral artery disease.

The incidence of PAD in Japan and South Korea is showing a downward trend, and the overall disease burden is lower than that in China and the Philippines. As major high-income countries in the Western Pacific region, both Japan and South Korea have a SDI at a high-middle level. In these two countries, higher levels of economic development, well-established healthcare systems, and strong public health awareness may collectively contribute to the reduction in the incidence and prevalence of PAD. Both nations are relatively well-equipped in terms of managing vascular disease risk factors, providing care for PAD patients, and allocating medical resources, thereby meeting the goals of universal health coverage. Additionally, surgical interventions related to PAD (such as vascular reconstruction techniques) and rehabilitation therapies are also more advanced in these countries. Notably, Japan’s low-salt diet culture and relatively low smoking rate play a positive role in reducing the onset and disability associated with vascular diseases[13,14]. Compared to high-income regions in Europe and North America, the PAD patient population in Japan and South Korea is more concentrated among the elderly. In high-income areas of Europe and North America, cases are primarily found among individuals aged 55 and above[1], whereas in Japan and South Korea, the affected populations are mainly those aged 70 and above and 60 and above, respectively. This difference may be related to the higher degree of population aging in the two Asian countries. As of 2021, Japan (with 28.9% of its population aged 65 or older[3]) and South Korea (with 16.5% of its population aged 65 or older[3]) exhibit significantly higher levels of population aging compared to high-income countries in Europe and North America. Given these epidemiological characteristics, Japan and South Korea should prioritize the elderly population and develop PAD management strategies tailored to the needs of an aging demographic structure.

For China and the Philippines, which have moderate SDI scores, the burden of PAD is relatively heavier. In recent years, China has seen a significant increase in both the prevalence and incidence of PAD, making it one of the countries with the highest number of PAD cases[15], and the disease burden is expected to further intensify in the future. Among the four countries included in this study, the Philippines faces the most severe PAD burden, with the highest age-standardized incidence rate (123.84 per 100,000), age-standardized prevalence rate (1,398.11 per 100,000), and age-standardized DALYs rate (11.82 per 100,000). The findings of this study are consistent with some previous research. Prior studies have found that adults in low- and middle-income regions are at significantly higher risk of developing PAD compared to those in high-income regions[16–18]. The results of this study suggest that both China and the Philippines are underinvesting in controlling PAD risk factors. Broader health resources and services are needed to meet medical demands. Both China and the Philippines still show notable shortcomings in achieving Universal Health Coverage (UHC). High-quality medical resources are concentrated in large cities, while remote and rural areas suffer from a lack of medical facilities. For example, major cities such as Manila in the Philippines and eastern regions of China are home to the majority of high-quality hospitals, whereas western China and Mindanao in the Philippines have insufficient medical resources, with some areas even facing shortages of doctors. Primary healthcare resources in both countries are inadequate, with low densities of primary care physicians and a small proportion of GDP allocated to healthcare expenditure, making it difficult to meet the needs for PAD prevention and treatment. Furthermore, existing studies have shown an overall negative correlation between the density of health workforce and vascular disease mortality and DALYs, indicating that increasing the human resources for vascular health should be a key priority[3,19,20]. The continuously worsening risk factors for non-communicable diseases also pose a serious threat to vascular health in both countries. For instance, China has a relatively high smoking rate[21]. Increased opportunities for both active smoking and exposure to secondhand smoke among men and women may be one of the main reasons for the growing PAD burden. Tobacco control should be one of the key measures to reduce the PAD burden. Health promotion programs and risk factor control initiatives are essential to preventing the increase of vascular diseases. Simple and cost-effective measures to reduce adverse outcomes mainly include the prevention and treatment of vascular disease risk factors, as well as regular monitoring for signs of lower limb disease in high-risk individuals and early referral to specialized services[11].

This study utilized reliable data from the GBD 2021 to report on the prevalence, incidence, and DALYs of PAD in two developed countries (Japan and South Korea) and two developing countries (China and the Philippines) in the Western Pacific region. Additionally, we projected the incidence of PAD in these four countries up to the year 2041. However, this study has several limitations. First, as with previous GBD studies, the GBD estimates rely on the quality and quantity of data sources available for informing the estimations. Data scarcity and poor quality in certain regions may lead to misestimation of disease burden, particularly in economically underdeveloped areas. Although the analytical processes and methodologies of the GBD have been continuously improved, the quality and collection of raw data remain major limiting factors. Therefore, this study did not include analysis of countries in the Western Pacific region with low sociodemographic development levels. Instead, it focused on four representative countries in the region with either high or medium sociodemographic development levels. Second, only a very small number of patients truly die directly from PAD. Patients may instead die from conditions such as myocardial infarction, stroke, or sepsis. As a result, reporting on PAD-related deaths is highly inaccurate, leading to an underestimation of the true burden of PAD.

Overall, there is considerable variation in the burden of PAD across countries with different SDI levels in the Western Pacific region. In higher SDI countries, the PAD burden has been gradually declining, whereas in lower SDI regions, the burden remains relatively heavy. PAD is expected to be an obstacle to future development in medium or low SDI areas. The burden of PAD also exhibits complex trends related to age, sex, and time, with particularly high burdens among the elderly. Efforts to prevent PAD have impacts far beyond reducing the incidence of PAD alone. They also yield positive effects on overall cardiovascular diseases.

## Data Availability

All relevant data are within the manuscript.The data are available from the Global Burden of Disease (GBD) study 2021 database(accession number Email?).

https://vizhub.healthdata.org/gbd-results/

## Notes

### Competing Interest Statement

The authors have declared no competing interest.

### Funding Statement

The author(s) received no specific funding for this work.

### Author Declarations

Only publicly available aggregated data from the Global Burden of Disease study were used, with no access to individually identifiable information. Therefore, ethical approval was not required for this study.

